# Influence of study characteristics, methodological rigor and publication bias on efficacy of pharmacotherapy in obsessive compulsive disorder: a systematic review and meta-analysis of randomized placebo-controlled trials

**DOI:** 10.1101/2023.11.24.23298989

**Authors:** S.E. Cohen, J.B. Zantvoord, B.W.C. Storosum, T.K. Mattila, J.G. Daams, B.N. Wezenberg, A. De Boer, D. Denys

## Abstract

**Question:** We examined the effect of study characteristics, risk of bias and publication bias on efficacy of pharmacotherapy in randomized controlled trials (RCT’s) for obsessive-compulsive disorder (OCD).

**Study selection and analysis:** We conducted a systematic search for double-blinded, placebo controlled short-term RCT’s with selective serotonergic reuptake inhibitors (SSRI’s) or clomipramine. We performed a random-effect meta-analysis, using change of the Yale-Brown Obsessive-Compulsive scale (YBOCS) as primary outcome. We performed meta-regression for key study characteristics, and for risk of bias. Furthermore, we analyzed publication bias using a Bayesian selection model.

**Findings:** We screened 3729 articles and included 21 studies, containing 4102 participants. Meta-analysis showed an effect size of −0.59 (Hedges’ G, 95% CI −0.73 to −0.46), equaling 4.2 point reduction on the YBOCS compared to placebo. The most recent trial was performed in 2007 and most trials were at risk of bias. In our meta-regression, we found that high risk of bias was associated with a larger effect size. Clomipramine was more effective than SSRI’s, even after correcting for risk of bias. We found an indication for publication bias subsequent correction for this bias resulted in a depleted effect size.

**Conclusions:** Our findings reveal superiority of clomipramine over SSRIs, even after adjusting for risk of bias. Effect sizes may be attenuated when considering publication bias and methodological rigor, emphasizing the importance of robust studies to guide clinical utility of OCD pharmacotherapy.

## Background

Obsessive-compulsive disorder (OCD) is characterized by persistent thoughts (obsessions) and repetitive behaviors (compulsions). Global life-time prevalence is two percent (1). Without treatment OCD may profoundly impair quality of life and social functioning. Selective serotonin reuptake inhibitors (SSRIs), cognitive behavioral therapy with exposure and response prevention (ERP), or a combination of both are currently recommended for management of OCD (2). Clomipramine was the first effective pharmacotherapeutic intervention, but its side-effect profile renders it a secondary option to SSRI’s (3).

Evidence for efficacy of SSRI’s in OCD and Major Depressive Disorder (MDD) is generally accepted and clinical use is common. However, quality and reporting of placebo-controlled RCT’s for MDD has recently been critically evaluated. Concerns have been raised regarding overestimation of SSRI efficacy in MDD due to flaws in trial quality (4–6). For instance, a recent reanalysis of a meta-analysis of randomized SSRI trials in MDD showed all studies to be of high or unclear risk of bias. Moreover, another meta-analysis only found a small, clinically irrelevant symptom reduction by SSRI’s (7, 8). While literature on quality and reporting of placebo-controlled RCTs in MDD has expanded in recent years, there is currently a paucity of data regarding quality of OCD trials, and the question whether overestimation of efficacy pervades in studies examining SSRIs’ in OCD remains currently unanswered.

Besides quality, study-level characteristics such as sponsor status, number of treatment arms and trial population have been found to influence trial success (9, 10). For OCD, there is currently a lack of information on the influence of RCT methodology on efficacy, even though this information could contribute in optimizing future trial design.

Recent evidence suggests that publication bias exaggerates pharmacotherapy efficacy in MDD, with more pronounced effect-size inflation in older trials (11). For OCD, a study examining publication bias compared efficacy measures of published trials with original FDA data. They found a pooled effect size of 0.39 Hedges’ G according to the US Food and Drug Administration (FDA), compared to 0.45 according to the published scientific literature. (12). This increase was non-significant, however this analysis only included SSRI trials submitted for FDA approval, potentially skewing the representation of reporting bias for OCD pharmacotherapy. Also, publication bias is often identified using funnel-plot models that focus on small-study effects, while selection models that focus on biases in publishing of significant studies might be more sensitive to publication bias. (13) (14). In addition, Bayesian approaches to selection models are especially suited for smaller meta-analyses. (15, 16).

## Objective

In this study, we examined the effect of study level characteristics on efficacy of pharmacotherapy in placebo-controlled OCD trials. Furthermore, we analyzed influence of risk of bias on effect size and investigated publication bias employing a Bayesian approach.

## Study selection and analysis

### Search strategy

We searched Embase, Medline and PsycINFO and Web of Science Conference Proceedings Index. Additionally, we searched the WHO International Clinical Trial Registry Platform search portal for registered studies, did a scoping search on Cochrane CENTRAL, and we used websites of several major conferences to search for unpublished literature or conference proceedings. We checked included articles for references and conducted citation screening. For a detailed account of our search strategy, see the supplementary material.

### Inclusion and exclusion criteria

Two investigators (S.E.C. and B.N.W.) included double-blind randomized-controlled trials for monotherapy with an SSRI or clomipramine in adult patients (18 years and older) with OCD. We included a non-selective patient population suffering from OCD with all subtypes. We included short-term studies with a primary end-point up to sixteen weeks using the Yale-Brown Obsessive-Compulsive Scale (Y-BOCS).

If studies did not publish quantified data or if we were unable to retrieve the full-text, we contacted the authors to request the information necessary for our analysis. If we were unable to retrieve this information, we excluded the study. Inclusion or exclusion conflicts were resolved by consensus or if necessary, through a consensus meeting with the co-authors. We pre-specified methods in the PROSPERO database for systematic reviews (registration number CRD42023394924).

### Data extraction

Two authors (S.E.C. and B.C.W.S.) extracted the following data from the included studies: mean age and gender, number of participants in active or placebo group, exclusion criteria, intervention and dosing regimen, washout period, time to primary endpoint and difference in YBOCS response for placebo and intervention. If a study used a fixed dosing regimen with multiple doses, we subdivided each dosing group and compared them to a proportionally reduced placebo group. Also, we extracted publication year, sponsor status and sponsor name, country or countries of trial site(s), use of a placebo run-in, number of trial site(s). To assess Risk of bias, we used the Cochrane RoB 2.0 tool (17). Risk of bias, subdivided into low, some concerns and high risk of bias, was assessed by S.E.C and B.C.W.S. Discrepancies were discussed in the entire research team in order to reach consensus.

### Meta-analytic method

As primary outcome, we used the mean change in YBOCS at primary study endpoint compared to baseline. If mean change scores were not reported, we used difference in YBOCS at study endpoint after ensuring baseline symptom severity to be balanced across intervention arms. For effect size, we used Hedge’s G, with a standard confidence interval (CI) of 95% using Knapp-Hartung adjustments (18, 19). Assuming between-study variability, we used a random-effects model for pooling effect sizes, with a restricted maximum likelihood estimator to calculate estimated SD of the true mean difference (τ) (20). We chose to pool clomipramine and SSRIs into one group, and carried out a meta-regression for potential moderating effect of intervention type (clomipramine or SSRI). Furthermore, we separately performed meta-regression for mean age, gender distribution, the use of two or more trial-arms, sponsor status, use of a placebo run-in phase, study size, risk of bias (high risk yes/no) and publication year. Since we use a multitude of single regressions, we adjusted the significance threshold by adjusting halfway p = 0.05 and the Bonferroni adjustment, i.e. p = 0.010. If moderators accounted for heterogeneity, we included them in a multiple meta-regression analysis. In order to avoid multi-collinearity, we tested for prediction correlators. If highly correlated (r>0.8), we included the moderator causing the highest amount of heterogeneity. For sponsorship status, we distinguished between studies that were fully sponsored by pharmacological companies, publicly funded studies, privately funded studies by any other institution than a pharmaceutical company and studies in which only medication was reimbursed by a pharmaceutical company. Additionally, we performed a separate meta-analysis for the SSRI group only, using the same methodology.

Effect of study quality was evaluated in a separate analysis by excluding studies that were at high risk of bias. For the moderating effect of trial quality, we included a sensitivity analysis excluding the randomization process from the risk of bias assessment, since randomization processes are currently defined more strictly compared to when the original studies were published (21).

### Publication bias

For publication bias analysis, we performed a fully Bayesian Copas Analysis using set, weakly informative, priors. Thus, we were be able to quantify the magnitude of publication bias in our selected studies using a measure D, and calculate the difference between corrected and non-corrected effect sizes, using a Bayesian meta-analysis. Additionally, we performed a corrected Egger’s test and used funnel plot for visual inspection of publication bias.

For meta-analytic estimation of pooled effect sizes, as well as for meta-regression and conventional publication bias quantification, we used the metafor and dmetar packages in R (22, 23). For Bayesian analysis for publication bias, we used the Robustbayesiancopas package in R (16).

## Findings

### Search results and study description

Our search yielded 3729 articles, of which we excluded 3646 after screening of the title and abstract. Of the 83 articles we included for full-text screening, we included twenty papers, one of which reported on two separate studies, making for a total of 21 studies (24). For a full account of our in- and exclusion, please refer to **figure 1** and the supplementary material. The 21 included studies contained a total of 4102 participants, with 739 participants in the clomipramine RCT’s and 3363 in the SSRI RCT’s. 51 percent of participants were male, 49 percent female. We included four studies focused exclusively on clomipramine, two of which were combined in a single manuscript. One three-armed study compared paroxetine to clomipramine and placebo. Of the included studies, all except four used a placebo run-in phase, most studies were fully sponsored by a pharmaceutical company (nine studies were supported by a grant independent from the producing company and of these eight, most were granted free use of medication). Nine studies used a fixed-dose regimen, one of which was a clomipramine study. For a full overview of study characteristics, see **table 1** and supplementary material.

**Figure 1.**
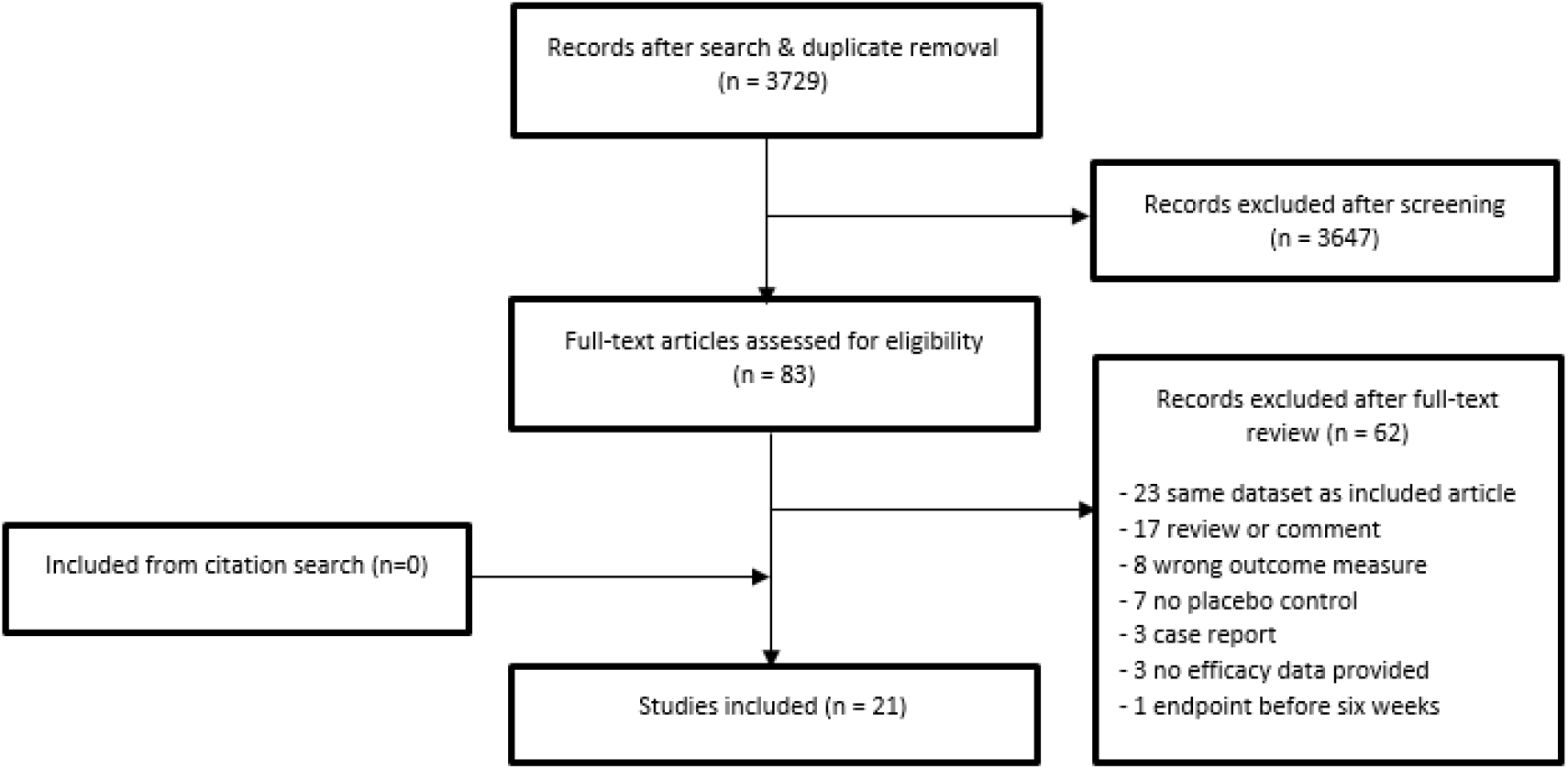
**F**low-chart of inclusion and exclusion of studies.

**Table 1:** study characteristics.

| Study | Drugs | Dose | N intervention/<br>placebo | Gender<br>M/F | Mean<br>age | Baseline<br>YBOCS | Treatment<br>duration | Sponsor | Duration<br>of illness<br>(years) | Primary<br>outcome | Placebo<br>run-in |
| --- | --- | --- | --- | --- | --- | --- | --- | --- | --- | --- | --- |
| CSG 1, 1991 | Clomipramine | 100 – 300<br>Flexible dose | 118/120 | 93/125 | 35.4 | 26.2 | 10 weeks | Ciba-Geigy<br>Corp | 15 | YBOCS<br>week 10 | 2-4 w |
| CSG 2, 1992 | Clomipramine | 100 – 300<br>Flexible dose | 123/129 | 128/13<br>5 | 35.6 | 26.7 | 10 weeks | Ciba-Geigy<br>Corp | 16, 3 | YBOCS<br>week 10 | 2-4 w |
| Chouinard,<br>1990 | Sertraline | 50-200 mg<br>dose-finding | 43/44 | 74/13 | 43.5 | 23 | 8 weeks | Pfizer | - | YBOCS<br>week 8 | 1 w |
| Foa, 2005 | Clomipramine | 200 – 250 mg | 36 / 26 | 34 / 28 | 34.8 | 23 | 12 weeks | NIMH | 16 | YBOCS<br>week 12 | - |
| Goodman,<br>1996 | Fluvoxamine | 100-300 mg | 78/78 | 78/78 | 36.7 | 23.25 | 10 weeks | Solvay | 16.5 | YBOCS<br>week 10 | 2-6 w |
| Goodman,<br>1989 | Fluvoxamine | 100 – 300 mg | 21/21 | 19/23 | 37 | 25.3 | 6 weeks | NIMH,<br>drugs by<br>Solvay | 15 | YBOCS<br>week 6 | - |
| Greist, 1995 | Sertraline | 50-100-200<br>mg | 79-81-80 / 84 | 191 /<br>133 | 38.7 | 23.7 | 12 weeks | Pfizer | 5.25 | YBOCS<br>week 12 | 1 w |
| Hollander,<br>2003 p | Paroxetine | 20-40-60 mg | 88 – 86 – 85 /<br>89 | 256 /<br>92 | 41.3 | 25.5 | 12 weeks | Smith-Kline<br>Beecham | - | YBOCS<br>week 12 | 2 w |
| Hollander,<br>2003 f | Fluvoxamine | 100 – 300 mg<br>flexible | 127 / 126 | 92 /<br>161 | 37.4 | 26.4 | 12 weeks | Solvay | 16.4 | YBOCS<br>week 12 | 1 w |
| Jenike, 1989 | Clomipramine | 200 – 300 mg<br>flexible | 13 / 14 | 13 / 14 | 39.4 | 25.7 | 10 weeks | Ciba-Geigy<br>Corp | - | YBOCS<br>week 10 | 2 w |
| Jenike, 1997 | Fluoxetine | 80 mg fixed | 23 / 21 | 23 / 21 | 35 | 19 | 10 weeks | NIMH grant<br>, Research<br>Fund | - | YBOCS<br>week 10 | 2 w |
| Jenike, 1990 f | Fluvoxamine | 50 – 300 mg<br>flexible | 18 / 20 | 20 / 18 | 36 | 22.7 | 10 weeks | Partly by<br>Solvay | 18.5 | YBOCS<br>week 10 | 2 w |
| Kamijima,<br>2004 | Paroxetine | 40 – 60 mg<br>flexible | 94 / 94 | 71 /<br>117 | 38 | 23.9 | 12 weeks | GSK | 10.5 | YBOCS<br>week 12 | 1 w |
| Kronig, 1999 | Sertraline | 50 – 200 mg<br>flexible | 86 / 81 | 92 / 75 | 36.5 | 25.2 | 12 weeks | Pfizer | 17.0 | YBOCS<br>week 12 | 1 w |
| Mallya, 1992 | Fluvoxamine | 50 – 300<br>flexible | 14 / 14 | 14/14 | 38 | 21.15 | 10 weeks | Not<br>sponsored | - | YBOCS<br>week 12 | 2 w |
| Montgomery<br>2001 | Citalopram | 20, 40, 60 mg<br>fixed | 102, 98, 100 /<br>101 | 184 /<br>217 | 37.8 | 25.6 | 12 weeks | Lundbeck | 15.9 | YBOCS<br>week 12 | 1 w |
| Montgomery,<br>1993 | Fluoxetine | 20, 40, 60 mg<br>fixed | 52, 52, 54 /<br>56 | 114 /<br>100 | 37 | 23.9 | 8 weeks | Lilly | - | YBOCS<br>week 8 | 1 w |
| Nakatani,<br>2005 | Fluvoxamine | 100 – 200 mg<br>flexible | 10 / 8 | 6 / 12 | 34.24 | 29.3 | 12 weeks | Gov.<br>research<br>grant | - | YBOCS<br>week 12 | - |
| Stein, 2007 | Escitalopram,<br>paroxetine | Esc 10 – 20,<br>par 40 fixed | 113, 114 /<br>117 / 114 | 197 /<br>261 | 38 | 27.1 | 25 weeks | Lundbeck | 4.1 | YBOCS<br>week 12 | - |
| Tollefson,<br>1994 | Fluoxetine | 20, 40, 60 mg<br>fixed | 87, 89, 90 /<br>89 | 159 /<br>196 | 37 | 22.7 | 12 weeks | Ely Lilly | - | YBOCS<br>week 12 | 1 w |
| Zohar, 1996 | Paroxetine,<br>clomipramine | 20-60 mg ,<br>50-250 mg<br>flexible | 201 / 99 / 99 | 209 /<br>209 | 38 | 26 | 12 weeks | SmithKline<br>Beecham | 15 | YBOCS<br>week 12 | 2 w |

Concise overview of included studies. N = number, M = man, F = female, YBOCS = Yale-Brown Obsessive-Compulsive Scale, w = week, CSG = Clomipramine Study Group, NIMH = National Institute of Mental Health, GSK = GlaxoSmithKline, p = paroxetine, f = fluvoxamine.

### Risk of bias

In total, four out of 21 included studies were judged as having a low risk of bias (see **figure 2**). Seven studies were at high risk of bias, and ten had some concerns. Concerns on randomization resulted mostly from absence of reporting on the allocation sequence generation process, and allocation concealment. Regarding assignment of intervention, four studies were at high risk of bias for not performing an intent-to-treat analysis, with drop-outs possibly leading to attrition bias. Four studies excluded more than ten percent of patients from the efficacy analysis. Risk of bias in measurement of outcome was low in all studies. In reporting of results, all but three studies did not report on the use a pre-specified analysis plan. For a full account of the risk of bias assessment, see the supplementary material. Our pre-defined sensitivity analysis excluding the randomization process from the risk of bias assessment did not change overall bias results.

**Figure 2.**
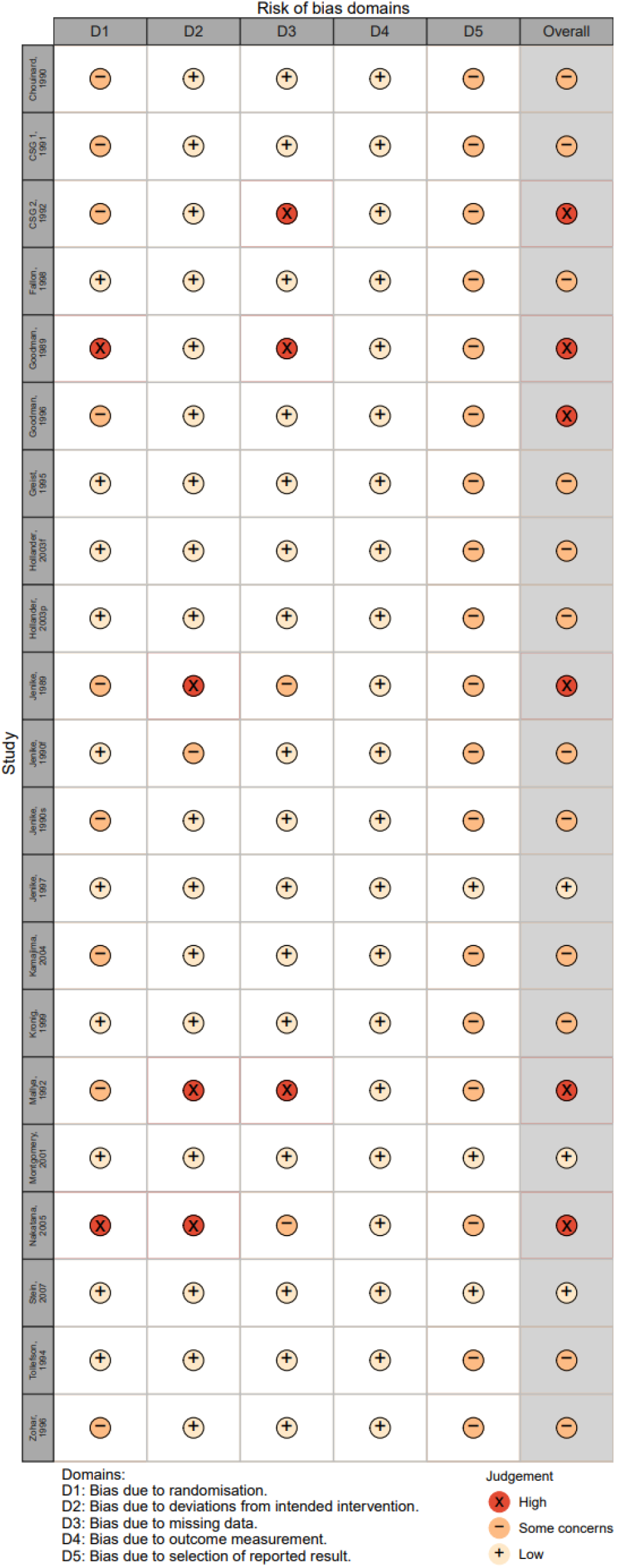
Risk of bias assessment for all studies, using the Cochrane Risk of Bias tool 2.0.

### General meta-analysis

A random effects meta-analysis with Knapp-Hartung adjustments resulted in a standardized mean difference (SMD) of - 0.59 (Hedges’ G, 95% CI −0.73 to −0.46, see **figure 3**), which is equal to a mean difference of 4.2 points on the YBOCS scale. The test for heterogeneity demonstrated a significant level of variability across the samples (Q = 114, p < 0.0001), with an estimated tau of 0.30, suggesting considerable between-study variance, and an I-squared value of 73%, indicating a high proportion of the observed variance reflects real differences in effect sizes. Heterogeneity was further demonstrated by the 95% prediction interval, which ranged from −1.22 to 0.03.

**Figure 3.**
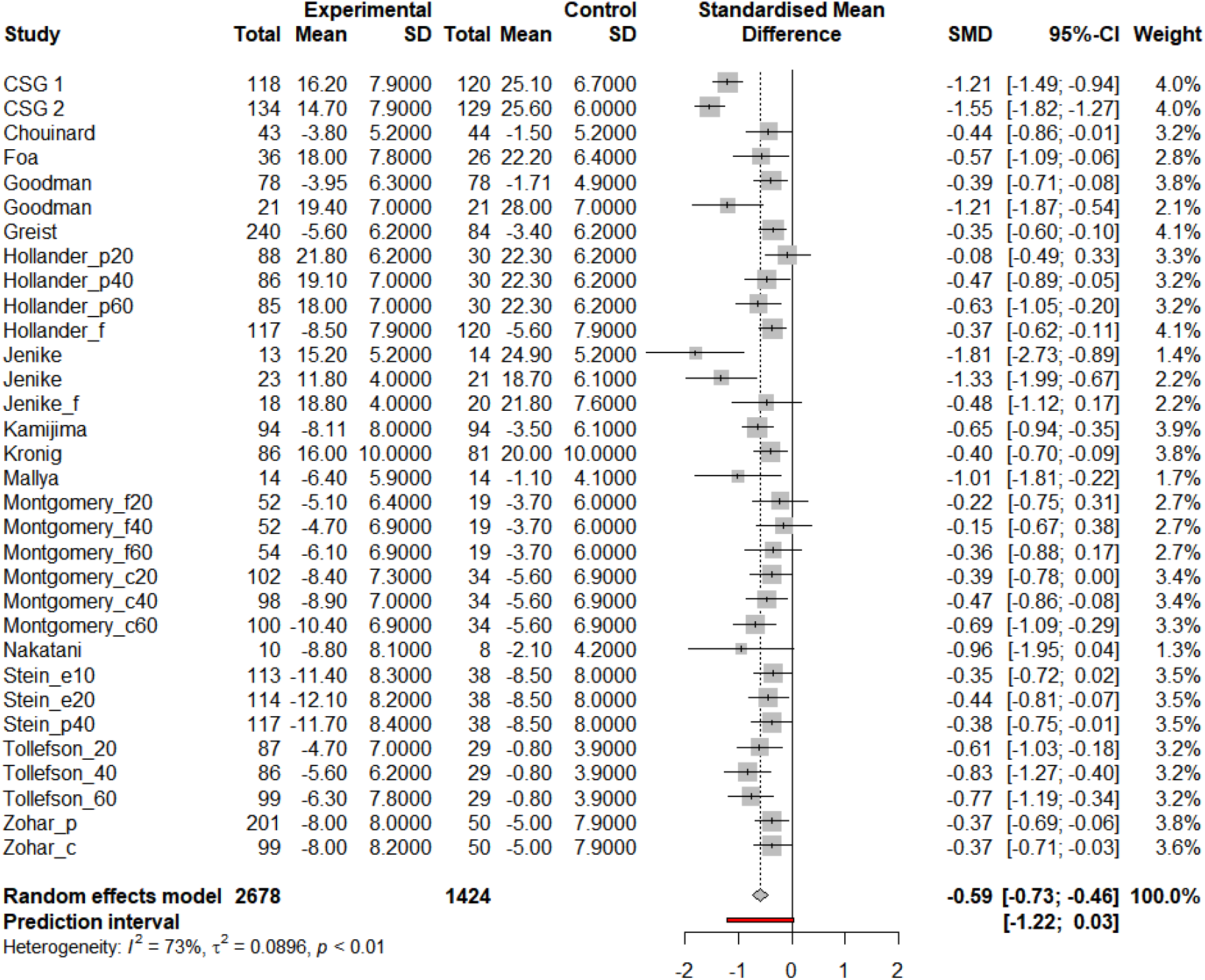
Forest plot of random effects meta-analysis of included studies.

### Meta-regression

After controlling for baseline differences, studies using clomipramine had a higher efficacy than SSRI studies (β = −0.55, 95% CI −0.24 to −0.9686 p = 0.0011), as were studies at high risk of bias (β = −0.51, 95% CI −0.19 to −0.82), p = 0.0029). For SSRI studies only, effect size was small (0.47 SMD, 95% CI - 0.56 to −0.39, equal to a YBOCS reduction of 3.5 points, see figure **S2** in the supplementary material for forest plot). Publication year was negatively associated with YBOCS change (suggesting that RCT efficacy decreases over time, β = 0.028, 95% CI = 0.0053 – 0.050, p = 0.017). Furthermore, studies that were fully sponsored by a pharmaceutical company had a smaller effect size than non- or partially sponsored studies (β = 0.46, 95% CI = 0.092 – 083, p = 0.016). Studies using two intervention arms had a higher efficacy compared to studies using one intervention arm (β = −0.33, 95% CI = - 0.061 – 0.60, p = 0.016). Using our corrected significance threshold (p = 0.010), publication year, sponsor status and amount of study arms did not reach significance level. We performed a separate meta-regression in which dosing arms of fixed-dose studies were combined in one intervention arm, which led to a non-significant influence of the amount of study arms on efficacy (see supplementary material). No other regression results differed in these sensitivity analyses. On participant level, increased mean age at baseline was associated with a decrease in efficacy (β = 0.075, 95% CI 0.0074 to 0.14, p = 0.031), which was insignificant after correcting for multiple analyses. Use of a placebo run-in, use of fixed or flexible dose, illness duration at baseline, baseline YBOCS and gender were not associated with changes in efficacy (See **table 2** for results of single meta-regression models).

**Table 2:**
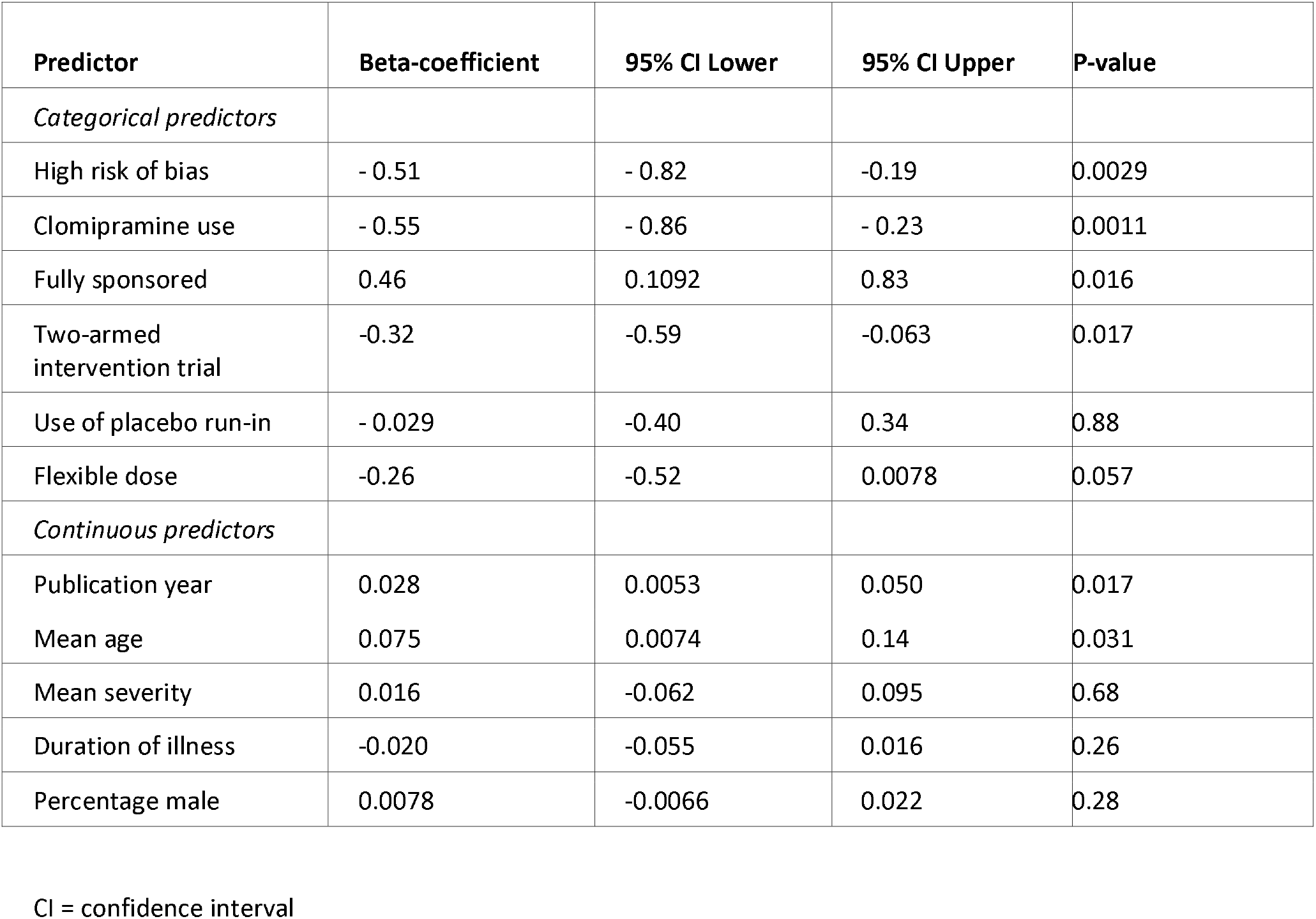
regression coefficients of single regressions.

| Predictor | Beta-coefficient | 95% CI Lower | 95% CI Upper | P-value |
| --- | --- | --- | --- | --- |
| *Categorical predictors* |  |  |  |  |
| High risk of bias | - 0.51 | - 0.82 | -0.19 | 0.0029 |
| Clomipramine use | - 0.55 | - 0.86 | - 0.23 | 0.0011 |
| Fully sponsored | 0.46 | 0.1092 | 0.83 | 0.016 |
| Two-armed intervention trial | -0.32 | -0.59 | -0.063 | 0.017 |
| Use of placebo run-in | - 0.029 | -0.40 | 0.34 | 0.88 |
| Flexible dose | -0.26 | -0.52 | 0.0078 | 0.057 |
| *Continuous predictors* |  |  |  |  |
| Publication year | 0.028 | 0.0053 | 0.050 | 0.017 |
| Mean age | 0.075 | 0.0074 | 0.14 | 0.031 |
| Mean severity | 0.016 | -0.062 | 0.095 | 0.68 |
| Duration of illness | -0.020 | -0.055 | 0.016 | 0.26 |
| Percentage male | 0.0078 | -0.0066 | 0.022 | 0.28 |

CI = confidence interval

Subsequently, we performed a multiple meta-regression analysis for significant predictors. Testing for multi-collinearity showed no redundant variables (see table **S2** in supplementary material for multi-collinearity table). We included risk of bias, clomipramine, sponsor status, number of intervention arms, mean age, publication year in our multiple meta-regression model using a mixed effect of maximum likelihood. Using our best performing meta-regression model, we found that when correcting for high risk of bias, clomipramine remained significantly correlated with a higher effect size compared to SSRI ( β = −0.43 with 95% CI −0.74 to −0.12, p 0.0085, **table 3**). For additional information regarding model performance and selection, see supplementary material.

**Table 3:** regression coefficients of multi-level meta-regression for the model including studies with high risk of bias and clomipramine use.

| Predictor | Beta-coefficient | 95% CI lower | 95% CI upper | p-Value |
| --- | --- | --- | --- | --- |
| High Risk of Bias | -0.33 | -0.64 | -0.0085 | 0.044 |
| Clomipramine Use | -0.43 | -0.74 | -0.12 | 0.0085 |

CI = confidence interval

### Publication bias

For the full sample, visual inspection of the funnel plot (**figure 4**), as well as the Egger’s linear regression test of funnel plot asymmetry, as implemented using the method by Pustejovsky, shows no indication of publication bias (t = 0.23, p-value = 0.82). In contrast, using a Bayesian Copas selection model, a moderate amount of publication bias was found (D = 0.48). After adjusting for publication bias, efficacy was reduced with 0.11 SMD, from - 0.53 (95% credible interval −0.64 to - 0.42) to - 0.42 (95% credible interval −0.60 to −0.22) using Bayesian analysis for determining efficacy.

**Figure 4.**
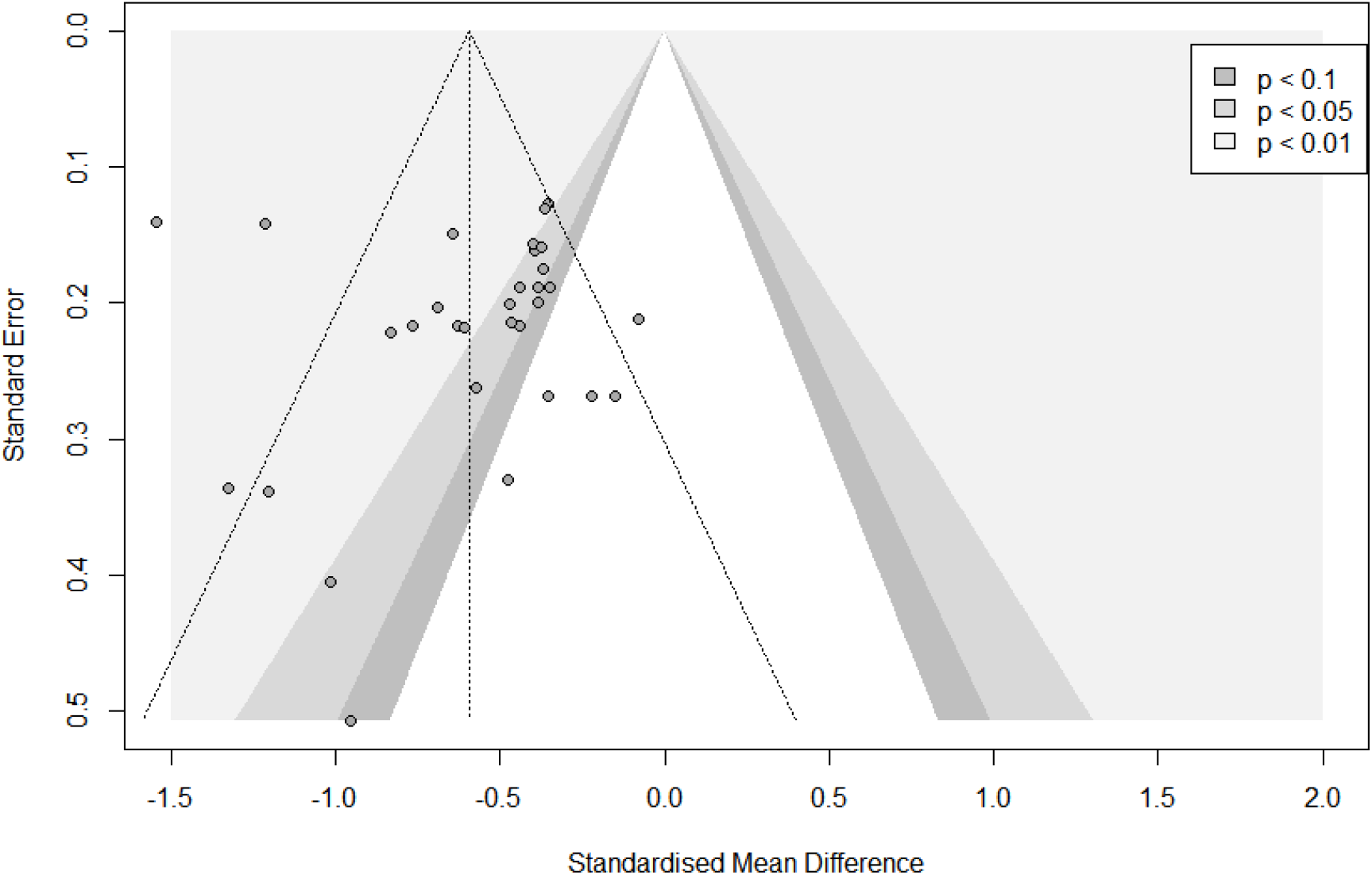
Funnel plot for visual inspection of publication bias.

## Conclusions and clinical implications

In our meta-analysis of RCT’s, we found that pharmacotherapy for OCD has a medium effect size in favor of intervention compared to placebo, amounting to 4.2 points on the YBOCS scale. Results were heterogeneous, with studies at high risk of bias being more likely to lead to a larger effect than studies that were at low risk of bias or had some concerns. Clomipramine was more efficacious than SSRI’s, even after correcting for studies at high risk of bias. Furthermore, using a Bayesian analysis we found a moderate risk of publication bias, with a small decrease in estimated efficacy after correcting for publication bias.

Our efficacy findings were comparable to those of an earlier meta-analysis and a recent network-meta-analysis (25, 26). The advantage of clomipramine over SSRI’s are also in line with earlier evidence (27). However, in contrast to our analysis, Skapinakis et al. found that clomipramine was not significantly more efficacious than SSRI’s and that any non-significant differences further dissipated when considering studies with low risk of bias. This result might be explained by the fact that the authors conducted a network analysis, also including head-to-head clinical trials. Furthermore, including only studies with low risk of bias is based on the stern assumption that studies with some methodological shortcomings do not have any added empirical value, and it depletes power for detecting subgroup analysis. Alternative explanations for clomipramine’s higher efficacy include lower quality and older studies conducted before SSRIs, suggesting the population was medication naïve with fewer non-responders. (27, 28). Although studies did not register the degree of earlier non-response, clomipramine remained a significant predictor for treatment success after correcting for publication year.

After correction for multiple tests, we did not find an effect of participant characteristics on trial efficacy, as is in line with earlier literature (29). Furthermore, study characteristics such as publication year, amount of intervention arms and study sponsorship were not significant after adjusting for multiple testing. Although a placebo-run in phase has been found to increase efficacy by decreasing placebo response in depression trials, we were unable to demonstrate an effect of placebo-run in, possibly as almost all studies actually used a run-in phase (30). Notably, the European Medicine Agency Guideline on clinical investigation of medicinal products for the treatment of OCD recommends against using a run-in phase as it might impair generalization of the study results (31).

We found only four studies with a low risk of bias, and seven studies with a high risk. The remainder of the studies were deemed at some concern for risk of bias. Most studies did not have a predefined analysis plan or protocol, and most studies did not disclose the randomization process or allocation procedure. Both might be explained by shifting publication and quality standards for RCT’s. More specifically, because almost all included studies were published before the 2004 requirement by journals for prospective registration in a public trial registry in and before the Food and Drug Administration mandated pre-registration in 2007 (32, 33). In fact, the most recent article we were able to include was published in 2007. In addition to critically appraising current evidence, we took into account the influence of risk of bias on effect size in OCD pharmacotherapy. To our knowledge, this is the first meta-analysis to do so, and we found that studies at high risk of bias are correlated with an increased effect size. This finding raises concerns regarding effect size inflation in pivotal OCD pharmacotherapy trials and emphasizes the need for (novel) high quality evidence on SSRI and clomipramine efficacy in OCD.

Our meta-analysis is the first study on OCD pharmacotherapy to demonstrate an effect of publication bias on efficacy, using a Copas selection model with a Bayesian approach. Selection models, which assume that publication bias arises from selective publishing of statistically significant studies, are thought to be preferable over funnel-plot methods for detection of publication bias (13, 34, 35). We chose to use a novel Bayesian selection model, as this approach circumvents the assumption of a normal distribution of effect sizes and is thought to enhance sensitivity in smaller meta-analyses (16). Not taking into account publication bias means potentially exaggerating treatment benefits which is highly relevant in clinical decision making. In the case of OCD, we conclude that, although there is clear evidence of publication bias in the literature and treatment benefit is diminished somewhat after correction, this effect is limited.

### Limitations

Our meta-analysis does have some limitations. Although we did perform meta-regression for participant-level characteristics such as age, gender and illness severity, using aggregate data meta-analysis for these interactions is problematic, since results might be falsely positive due to the ecological fallacy, in which treatment effects are confounded by a third, unknown factor (36). On the other hand, potential modifiers on the participant level might also be missed if studies did not show variation in aggregate measures. Specifically for inspection of patient-level modifiers, the designated research method would be an individual participant data meta-analysis (IPDMA), which to the best of our knowledge has not been performed yet, specifically for OCD pharmacotherapy (37) Finally, as we only included placebo-controlled trials, our results pertain to a clinical trial population which might decrease representativeness for real-world population regarding for instance co-morbidity and treatment resistance (38).

### Clinical implications and recommendations for future research

Although our findings extend current literature by supporting evidence for the efficacy of OCD pharmacotherapy, the effect size is diminished after correcting for methodological issues and publication bias (27, 39). Furthermore, the fact that most placebo-controlled trials for OCD, based on registered medications, fall short on contemporary quality standards, is a concerning matter.

When interpreting these results, we should also differentiate between statistical and clinical relevance. It is questionable whether an active medication vs. placebo separation of 4.2 points on the YBOCS, and even less for SSRI’s, amounts to a noticeable change in the quality of life of a patient, after outweighing possible side-effects. The term “minimal important difference” (MID) describes the smallest change seen as beneficial by patients (40). No generally accepted MID exists for the YBOCS, and to our knowledge the most recent study proposing an MID used a five points difference, a value slightly above our findings (41).

The advantage in efficacy of clomipramine over SSRI’s persisted even after correcting for risk of bias, which raises questions regarding its secondary ranking in current prescription guidelines. However, direct head-to-head trials have not shown a clear increase in efficacy of clomipramine over SSRI’s, and clomipramine is known to have a more severe side effect profile compared to SSRI’s, especially anticholinergic, cardiac and metabolic side effects (28). As such, we are not able to recommend concrete changes in clinical practice, based on this article. Yet it is noteworthy that clomipramine, the inaugural approved pharmacological treatment for OCD in 1989, may remain one of our most potent treatment options.

Given that study effects are modest, susceptible to publication bias and influenced by the risk of bias, a new RCT reassessing the efficacy of OCD pharmacotherapy would hold significant clinical and regulatory importance. This trial should be conducted in a representative population, adhere to the latest quality standards, and prioritize social functioning and quality of life, alongside symptom reduction. However, we acknowledge the financial burdens associated with conducting such a RCT and the ethical concerns surrounding the inclusion of a placebo arm, thereby potentially depriving participants of an established first-line treatment for OCD (20). In light of these considerations, undertaking an individual IPDMA of OCD RCTs to first assess patient-level characteristics predicting improved treatment response would be both economically and ethically more favorable. Such subgroup analyses could refine interventions, identifying specific patient populations with enhanced treatment success likelihoods. Consequently, insights from an IPDMA could inform targeted cohorts for upcoming drug trials.

In summary, our findings suggest a pronounced efficacy for clomipramine over SSRIs, even after adjusting for risk of bias. While pharmacotherapy remains a viable therapeutic option for OCD, it’s essential to recognize that reported effect sizes may be attenuated when considering publication bias and stringent methodological standards.

## Data Availability

All data produced in the present study are available upon reasonable request to the authors

