## Supplementary material for "Influence of study characteristics, methodological rigor and publication bias on efficacy of pharmacotherapy in obsessive compulsive disorder: a systematic review and meta-analysis of randomized placebo-controlled trials"

*Search strategy*

Using a sensitivity-maximizing search, we included terms regarding the population and intervention, using an RCT filter. We did not use language or date restrictions. The search was conducted by author JD, clinical librarian and search specialist, in order to ensure a high degree of thoroughness. For population /domain being studied, we included the term obsessive-compulsive disorder and known synonyms. For intervention we used pharmacotherapy for OCD as recommended in the NICE treatment guideline and in the Anxiety and Depression Association of America treatment guideline. We searched for clomipramine, sertraline, paroxetine, fluoxetine and fluvoxamine, incuding known synonyms. Citalopram, escitalopram, mirtazapine and venlafaxine are not registered but are mentioned as treatment options in abovementioned guidelines, so we included them in our search.

We systematically searched Embase, Medline and PsycINFO. For the Embase search strategy, see figure **S1**. Comparable searches were done for Medline and PsycINFO**.** Additionally, we performed a scoping search of Cochrane CENTRAL which did not yield additional articles. We searched the WHI International Clinical Trial Registry Platform, as well as EUdraCT and clinicaltrials.gov. Additionally, we searched several symposia (ACNP, ECNP, Molecular Psychiatry, ADAA, IOCDF) for the last five years in order to included information that has not yet been published.

**Figure S1: Embase search strategy**
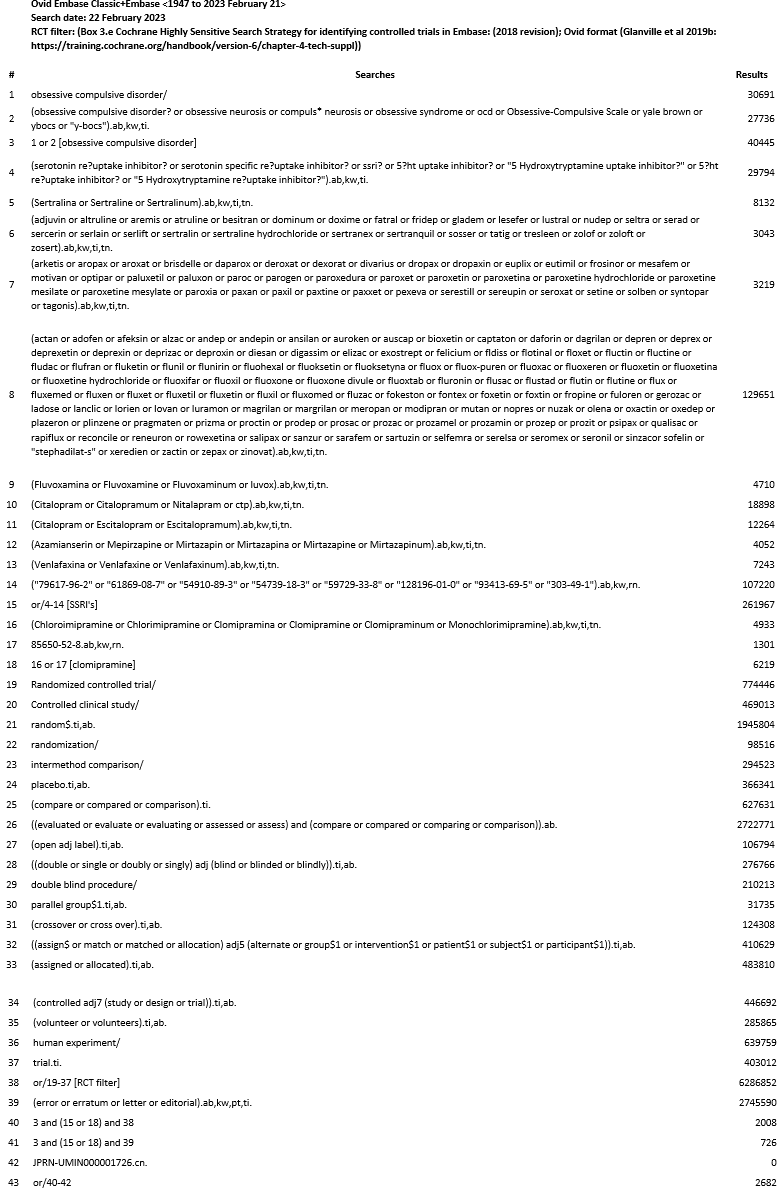

*In- and exclusion of studies.*

With full-text screening, we excluded 23 studies for using the same data from a trial that had already been presented in an earlier paper. We excluded eight studies for not using the YBOCS scale as outcome measure and seven for not using a placebo control group. 17 were excluded because they were a review or comment and three for presenting a case report. Three papers did not provide enough efficacy data to include them in our review, even after requests for information. We excluded one study for using 24 hours as endpoint, after administering intravenous clomipramine.

*Risk of bias assessment.*

We used the [Cochrane Risk of Bias 2.0 tool](https://methods.cochrane.org/risk-bias-2), and through the official guidance document we filled in the risk of bias template for each study. See **table S1** , in which we simplified and summarized our risk of bias assessment.

**Table S1: Risk of bias assessment**

| **Study** | **Domain 1**  **Randomization** | **Domain 2a**  **Assignment** | **Domain 3**  **Missing outcome data** | **Domain 4**  **Outcome measurement** | **Domain 5**  **Reporting** | **Overall risk of bias** |
| --- | --- | --- | --- | --- | --- | --- |
| Chouinard, 1990 | Some concerns  1.1./1.2 : Unclear allocation sequence / concealment | Low | Low | Low | Some concerns  5.1 No pre-specified analysis plan. | Some concerns |
| CSG 1, 1991 | Some concerns  1.1./1.2 : Unclear allocation sequence / concealment | Low | Low | Low | Some concerns  5.1 No pre-specified analysis plan. | Some concerns |
| CSG 2, 1992 | Some concerns  1.1./1.2 : Unclear allocation sequence / concealment | Low | 3.1 not all ptcpts that were randomized, were analyzed | Low | Some concerns  5.1 No pre-specified analysis plan. | High |
| Goodman, 1989 | Some concerns  1.1./1.2 : Unclear allocation sequence / concealment | Low | High  3.1 No ITT analysis, >10 % dropout | Low | Some concerns  5.1 No pre-specified analysis plan. | High |
| Goodman, 1996 | High. 1.1/1.2 unclear, and  1.3 Allocation, age and gender all identical. Exceeds chance expecation | Low | Low | Low | Some concerns  5.1 No pre-specified analysis plan. | High |
| Greist, 1995 | Low | Low | Low | Low | Some concerns  5.1 No pre-specified analysis plan. | Some concerns |
| Hollander, 2003f | Low | Low | Low | Low | Some concerns  5.1 No pre-specified analysis plan. | Some concerns |
| Hollander, 2003p | Low | Low | Low | Low | Some concerns  5.1 No pre-specified analysis plan. | Some concerns |
| Jenike, 1989 | Some concerns  1.1./1.2 : Unclear allocation sequence / concealment | High  2.6/2.7 ITT unclear possible impact on results | Some concerns  3.1 ITT unclear, >10% dropout | Low | Some concerns  5.1 No pre-specified analysis plan. | High |
| Jenike, 1990f | Low | Some concerns  2.6 no ITT, 2 dropouts, minor | Low | Low | Some concerns  5.1 No pre-specified analysis plan. | Some concerns |
| Jenike, 1997 | Low | Low | Low | Low | Low | Low |
| Kamajima, 2004 | Some concerns  1.1./1.2 : Unclear allocation sequence / concealment | Low | Low | Low | Some concerns  5.1 No pre-specified analysis plan. | Some concerns |
| Kronig, 1999 | Low | Low | Low | Low | Some concerns  5.1 No pre-specified analysis plan. | Some concerns |
| Mallya, 1992 | Some concerns  1.1./1.2 : Unclear allocation sequence / concealment | High  2.6/2.7 No ITT analysis, >10 % attrition | High  3.1 No ITT analysis, >10 % dropout | Low | Some concerns  5.1 No pre-specified analysis plan. | High |
| Montgomery, 2001 | Low | Low | Low | Low | Low | Low |
| Montgomery, 1993 | Low | Low | Low | Low | Low | Low |
| Nakatana, 2005 | High  1.1 allocation not random | High  2.6/2.7 No ITT analysis, >10 % attrition | Some concerns  3.1 >10 % dropout | Low | Some concerns  5.1 No pre-specified analysis plan. | High |
| Stein, 2007 | Low | Low | Low | Low | Low | Low |
| Tollefson, 1994 | Low | Low | Low | Low | Some concerns  5.1 No pre-specified analysis plan. | Some concerns |
| Zohar, 1996 | Some concerns  1.1./1.2 : Unclear allocation sequence / concealment | Low | Low | Low | Some concerns  5.1 No pre-specified analysis plan. | Some concerns |

*Meta-regression analysis*

For studies included in our multiple metaregression, we used a multicollinearity test in order to avoid overfitting, whereby studies with a high correlation (r>0.8) would be excluded from the multiple meta-regression. As **table S2** shows, no studies were correlated to the degree of redundancy.

**Table S2: multicollinearity testing**

|  | **Publication Year** | **Trial arms** | **Sponsor status** | **High Risk of Bias** | **Clomipramine use** |
| --- | --- | --- | --- | --- | --- |
| Publication Year |  | -0.37 | 0.22 | -0.22 | -0.24 |
| Trial arms | -0.37 |  | -0.37 | -0.53 | -0.20 |
| Sponsor status | 0.22 | -0.37 |  | -0.63 | -0.24 |
| High Risk of Bias | -0.22 | 0.53 | -0.63 |  | 0.40 |
| Clomipramine Use | -0.24 | -0.20 | -0.19 | 0.40 |  |

Using anova, we compared performance and correctness of fit of the different multiple meta-regression models. The multiple metaregression using clomipramine and high risk of bias performed significantly better than individual regression models (see table **S3**). Further increasing model complexity did not lead to a significantly better performance. Corrected Akaike’s information criterion was lowest for the model using clomipramine and high risk of bias (see table **S4**). Using the parsimony principle, the metaregression with high risk of bias and clomipramine was preferred over more complex models. Notable, furthermore, is that even when using the most complex model including all metaregression variables, clomipramine remained a significant predictor (beta -0.39, 95%CI -0.70 to -0.076, p = 0.017).

**Table S3: Model performance of metaregressions**

| **Comparison of model performance** | **LRT** | **p-value** |
| --- | --- | --- |
| Clomipramine + High RoB vs. High RoB | 6.9 | 0.009 |
| Clomipramine + High RoB vs. Clomipramine | 4.9 | 0.0276 |
| Clomi + High RoB + sponsor status vs Clomipramine + High RoB | 1.5 | 0.22 |
| Clomipramine vs. High RoB vs Clomipramine + High RoB + publication year | 2.8 | 0.10 |
| Clomi + High RoB + sponsor status vs Clomi + High RoB + sponsor status + publication year | 2.7 | 0.10 |
| Clomi + High RoB + sponsor status vs. full model | 4.2 | 0.12 |

LRT: likelihood ratio test statistic

**Table S4: Akaike’s information criterion of metaregressions**

| Variables in regression model | AICc |
| --- | --- |
| High risk of bias | 26.1 |
| Clomipramine use | 24.1 |
| High risk + clomipramine | 21.8 |
| High risk + clomipramine + publication year | 21.8 |
| High risk + clomipramine + sponsor status | 23.2 |
| Full model | 23.6 |

AICc = Corrected Akaike’s information criterion

*Meta-analysis of SSRI studies*

Random effects meta-analysis of RCT’s for SSRI’s, resulted in a small effect size of -0.47 SMD (Hedges g, 95% CI -0.56 to -0.39, see **figure S2**). Heterogeneity was low across studies (I squared = 16.0%, tau < 0.0001), and the test for heterogeneity was not significant (Q = 30, p = 0.23), suggesting the effect of SSRIs compared to placebo to be consistent across studies. Meta-regressions for different SSRI’s were not significant. Results persisted when considering a prediction interval (95% PI -0.55 to -0.39).

**Figure S2** Forest plot for SSRI studies only

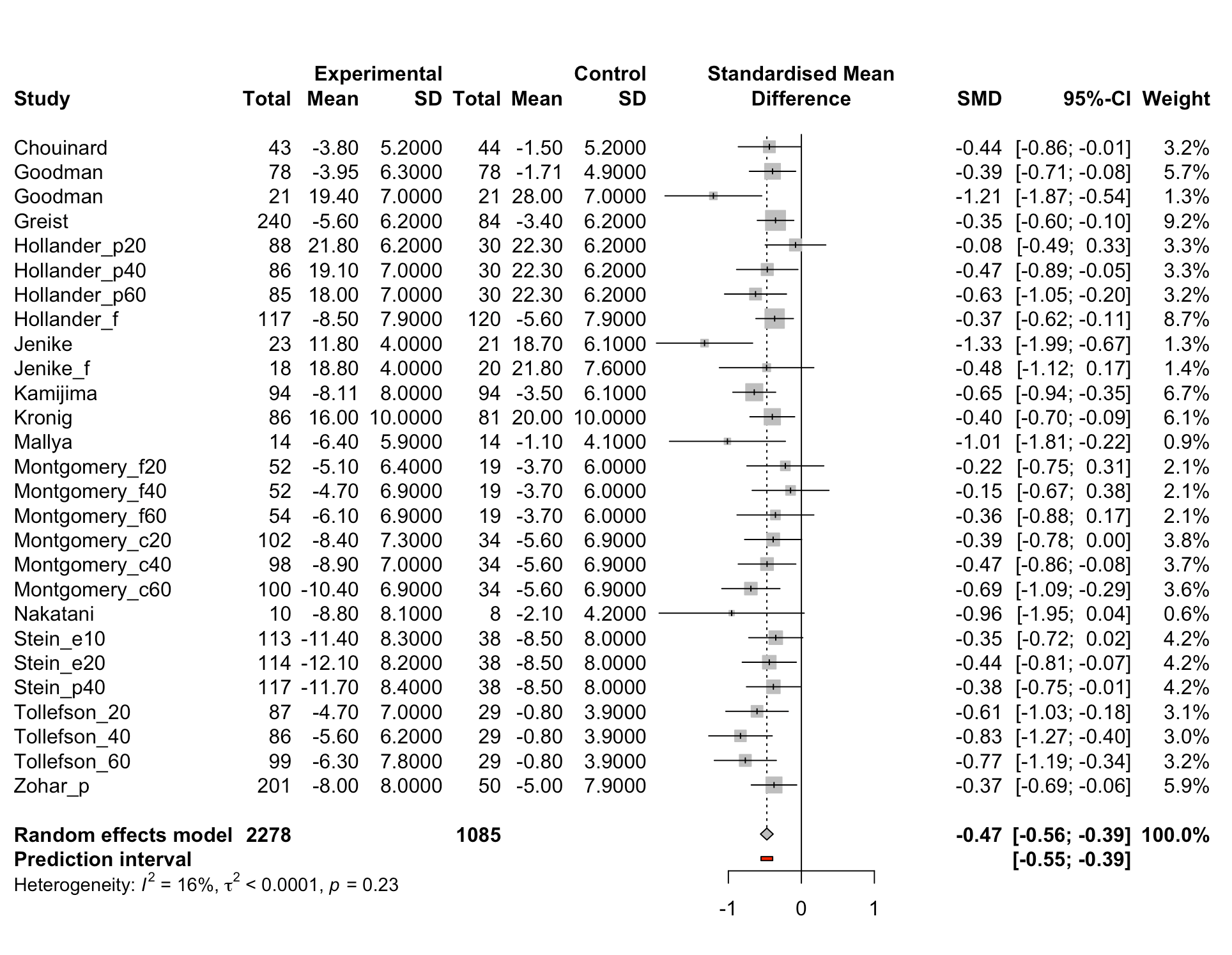

*Publication bias*

We used the robustbayesiancopas package in order to perform our Bayesian analysis of selection bias and used their proposed methods. We used multiple assumptions about distribution of the random effect (Student’s T, Laplace, normal and slash distributions). Then, we extracted the Deviance Information Criterion (DIC) for each model to compare their goodness of fit. As slash distributions had the best fit (i.e. the lowest DIC), we used this distribution in further calculations., We then estimated the correlations parameter and fit a Bayesian model with and without correction for bias. We repeated our analysis multiple times using different seed settings which did not change the results. For SSRI studies only, using a Bayesian Copas selection model, we found a moderate effect of publication bias (D = 0.48) similarly to the full sample, with a decrease of 0.077 SMD, from – 0.48 (95% credible interval -0.57 to -0.40) to – 0.41 (95% credible interval -0.54 to -0.22).

*Sensitivity analysis*

After fully excluding all studies with a high risk of bias, clomipramine was still associated with a higher effect size (-0.38, p = 0.028, 95% CI =-0.72 to - 0.044), emphasizing the robustness of our finding that clomipramine has a higher efficacy than SSRI’s when compared to placebo.

After combining intervention arms using different fixed doses, efficacy measures were comparable (SMD = -0.65, 95% CI -0.83 to -0.46). See **figure S3** for forest plot, including measures of heterogeneity. Furthermore, outcomes of meta-regression remained largely unchanged, except non-significance of the amount of intervention arms that were used. As our original analysis method increases the relative weight of studies with multiple intervention arms, the fact that in this analysis intervention arms are not significantly related to efficacy is an important addition to our original fidings. Please see **table S5** for single meta-regression results, and **table S6** for multiple metaregression including high risk of bias and clomipramine use. Furthermore, precision of estimates broadly decreased, with higher p-values, which is understandable considering the combination of doses decreases the degree of freedom for meta-regressions.

**Figure S3** forest plot of studies with fixed doses combined in a single intervention arm.

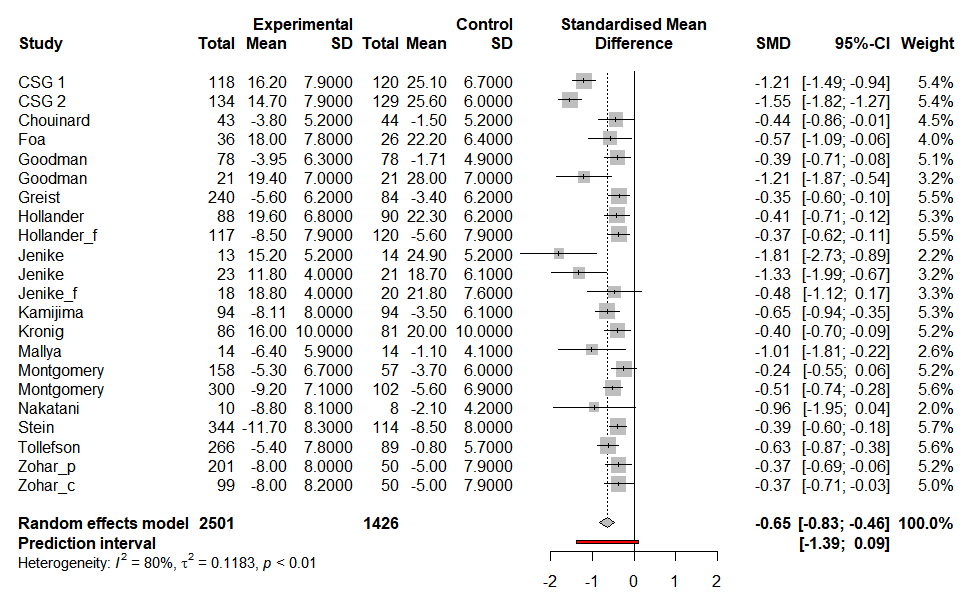

**Table S5** single regression outcomes for combined fixed doses.

| **Predictor** | **Beta-coefficient** | **P-value** | **95% CI Lower** | **95% CI Upper** |
| --- | --- | --- | --- | --- |
| *Categorical predictors* |  |  |  |  |
| High risk of bias | - 0.49 | 0.012 | - 0.87 | -0.12 |
| Clomipramine use | -0.53 | 0.0073 | -0.90 | -0.16 |
| Fully sponsored | 0.44 | 0.041 | 0.020 | 0.87 |
| Two-armed intervention trial | -0.33 | 0.060 | -0.67 | 0.015 |
| Use of placebo run-in | 0.052 | 0.84 | -0.47 | 0.58 |
| Flexible dose | -0.25 | 0.17 | -0.62 | 0.11 |
| *Continuous predictors* |  |  |  |  |
| Publication year | 0.031 | 0.048 | 0.0030 | 0.062 |
| Mean age | 0.074 | 0.091 | -0.013 | 0.16 |
| Mean severity | -0.0038 | 0.94 | -0.11 | 0.10 |
| Duration of illness | -0.019 | 0.44 | -0.072 | 0.034 |
| Percentage male | 0.0075 | 0.50 | -0.015 | 0.030 |

**Table S6** multiple regression outcomes for combined fixed doses

| **Predictor** | **Beta-coefficient** | **p-Value** | **95% CI lower** | **95% CI upper** |
| --- | --- | --- | --- | --- |
| High Risk of Bias | -0.34 | 0.069 | -0.70 | 0.029 |
| Clomipramine Use | -0.43 | 0.022 | -0.79 | -0.070 |
